# Study protocol for estimating modern US social contact patterns: the ENGAGED study

**DOI:** 10.64898/2026.01.08.26343704

**Authors:** Machi Shiiba, Marina Bruck, Musu M Sesay, Alexander F Hudgins, Marni F. Segall, Pragati Prasad, Charlotte R Doran, Aaron J Siegler, Felipe Lobelo, A. Blythe Ryerson, Ben Lopman, Kristin Nelson, Sharia M Ahmed

## Abstract

**Background:** Accurately capturing social contact data is essential for developing effective mathematical models to forecast disease trends and evaluate interventions. There are limited population-based data of social contacts in the United States (US) which limits our ability to accurately model infectious disease transmission.

**Methods:** To fill in this gap, we conducted a staggered longitudinal cohort study in metropolitan Atlanta, Georgia, USA. We aimed to characterize contact patterns and examine how they varied by i) participant demographics, ii) seasonality, and iii) self-managed and medically-attended symptoms. Once per month for six months, participants reported individual contacts they can name, individual contacts they cannot name, and contacts that occurred in group settings. We defined individual contacts as a two-way conversation with five or more words in the physical presence of another person or physical skin-to-skin contact, and group contacts as contacts with a group of people with whom participants talked, interacted, or shared space. Participants were enrolled on a rolling basis, and data is collected from November 2024 through April 2026. Data analysis will generate age-specific contact matrices using individual contacts and compare contact rates by symptoms. We will also analyze the number and characteristics (e.g. indoor/outdoor) of each type of contacts. The contact matrices and results will be publicly available for the wider modeling community.

**Discussion:** This study is among the first population-level longitudinal studies of social contact patterns in the US. By capturing behavioral changes during periods of both health and illness, and across seasons, this study will provide insights into the complex dynamics of human behavior relevant to infectious disease transmission in the US.

## Introduction

Lower respiratory and enteric infections constitute a significant burden of infectious disease morbidity worldwide and are major causes of mortality among children under 5 years of age (1, 2). Person-to-person interactions, or contacts, drive the transmission of such infections (3). Mathematical models are increasingly used to study disease dynamics, forecast disease trends, and evaluate interventions such as vaccines and school closure (4–6). One key to accurate mathematical models is correctly accounting for human behavior in disease transmission.

The POLYMOD study was an early effort to estimate social contact rates and was conducted in eight European countries from 2005 through 2006 (7). Social contact rates from POLYMOD have been widely used for infectious disease modeling studies, including in countries outside of Europe (8–13). However, social contact patterns vary by culture, demographics, and socioeconomic status (13–15), and we lack recent, generalizable estimates for the United States (US). Previous US studies focused on school settings, households, or business offices, which may not generalize to populations outside those contexts (16–21).

Furthermore, social contact studies generally focus on behavior when participants are healthy, or during the acute phases of the COVID-19 pandemic. However, people tend to change their social behaviors while ill. Several studies reported that people had fewer contacts during periods of symptomatic illness, largely driven by reduction in contacts outside the home such as in schools and workplaces, leading to different contact age distribution when people were ill (22, 23). Additionally, seasonality influences social contact patterns. Colder weather has been linked to an increase in the average number of contacts, especially longer duration contacts (24, 25). Incorporating such behavioral changes by health status and seasons into models is essential for improving predictions of disease transmission dynamics.

We are conducting a staggered longitudinal cohort, using monthly online surveys to collect self-reported social contact data from participants of all ages in the state of Georgia, US. By collecting longitudinal contact data and linking participants’ health data, we aimed to explore how contact patterns vary across health status, seasons, and demographic factors. This approach will contribute to more accurate modeling of infectious disease transmission by capturing dynamic and context-specific contact behavior.

## Materials and methods

### Study design and objectives

This is a staggered longitudinal cohort study, with each participant contributing up to 6 months of data, and a rolling enrollment process for a total of 18 months of data collection. The main objective of the study is to collect data on routine social contacts using repeated monthly social contact diaries on a representative sample of metropolitan Atlanta residents in the state of Georgia, US. Additionally, we aimed to characterize contact patterns i) by participant demographics, ii) by seasonality, and iii) by self-managed and medically-attended symptoms (sought care at Kaiser Permanente healthcare facilities, see Event-triggered contact survey, medically-attended symptoms section).

### Study methods

Study participants were recruited from Kaiser Permanente Georgia (KPGA), an integrated healthcare system serving the greater metropolitan area of Atlanta, Georgia. At the end of 2025, KPGA insured more than 330,000 members (63% employer-sponsored). Members are mostly adults 18−64 years of age (70%), female (56%), and are Black/African American (43%) or White (25%), which roughly reflects the demographics of the region.

### Inclusion & exclusion criteria

KPGA members of all ages were eligible to be included in the study if they had an active KPGA membership at time of data query for eligibility and had an email address on file. Potential participants were categorized based on their age – children (under 13 years of age), teens (aged 13-17 years), and adults (18 years and above). For children and teens under the age of 18 years, their designated caregiver(s) on their medical record must have had an email on file to be included in the study.

Individuals were excluded from the study if they or their responding caregivers did not provide consent or parental permission online, unable to complete the remotely self-directed study activities (e.g., provide online consent or complete online surveys) in English, or reported cognitive impairment. Potential participants were excluded from study outreach if their records on file indicated ‘do not contact’ for research purposes.

Participants who would turn 7 or 18 years old within 30 days of receiving an enrollment invitation were excluded from that specific recruitment wave to ensure accurate assignment to age-specific consent or assent forms and survey questionnaires. These individuals remained eligible in subsequent recruitment waves.

### Participant selection and sample size calculation

KPGA constructed a stratified random sample of their active membership based on a target demographic distribution once a month from November 2024 – November 2025 to identify eligible potential participants for recruitment outreach and enrollment. There was a three-month new participant recruitment pause from April – June 2025 due to volatility in federal funding streams. We continued to collect data from existing participants during this time. Enrollment began on November 14, 2024 and concluded December 11, 2025, with data collection to continue through April 2026.

Initially, the study aimed at inviting 15,200 KPGA members (potential participants) and assumed a 5% response rate to enroll 760 participants over a 12-month period with anticipated enrollment of a total of 560 adults, 100 teens, and 100 children. This sample size as well as age breakdown was determined to allow sufficient precision to identify differences in contact patterns between demographic groups and calendar month based on observed contact rates in a prior study among US adults (20). Members were recruited within strata of age, sex, and race/ethnicity proportional to the distributions matching the metro Atlanta area. Due to new budget constraints in early 2025, the target outreach and enrollment were reduced to 11,500 KPGA members invited and 690 participants. The revised enrollment targets assumed an increased response rate of 6% based on early participant enrollment numbers, and the reduced enrollment number of 70 participants from only the adult participant group.

### Regulatory compliance

Kaiser Permanente Interregional Institutional Review Board (KPiIRB), the IRB of record for this study, reviewed and approved all submitted study documents - study protocol, survey and recruitment materials. Following IRB study approval, an IRB Authorization Agreement (IAA) was established with the Emory University IRB documenting the roles and responsibilities of each study site. KPGA established a data use agreement (DUA) with Emory University outlining the data use terms, including study variables and de-identified datasets to be transferred to the Emory team and the data transfer cadence and frequency. All study team members have access to a copy of the study protocol and DUA for reference to ensure adherence to approved study procedures.

### Invitation, screener, and consent

All study communication occurred through a secure REDCap portal hosted within KPGA. Potential participants received a recruitment email invitation containing instructions and an online link to the secure REDCap portal to review study information and complete enrollment. Non-respondents received weekly recruitment email invitations for 2 consecutive weeks after the initial recruitment email invitation (Figure 1).

**Figure 1:**
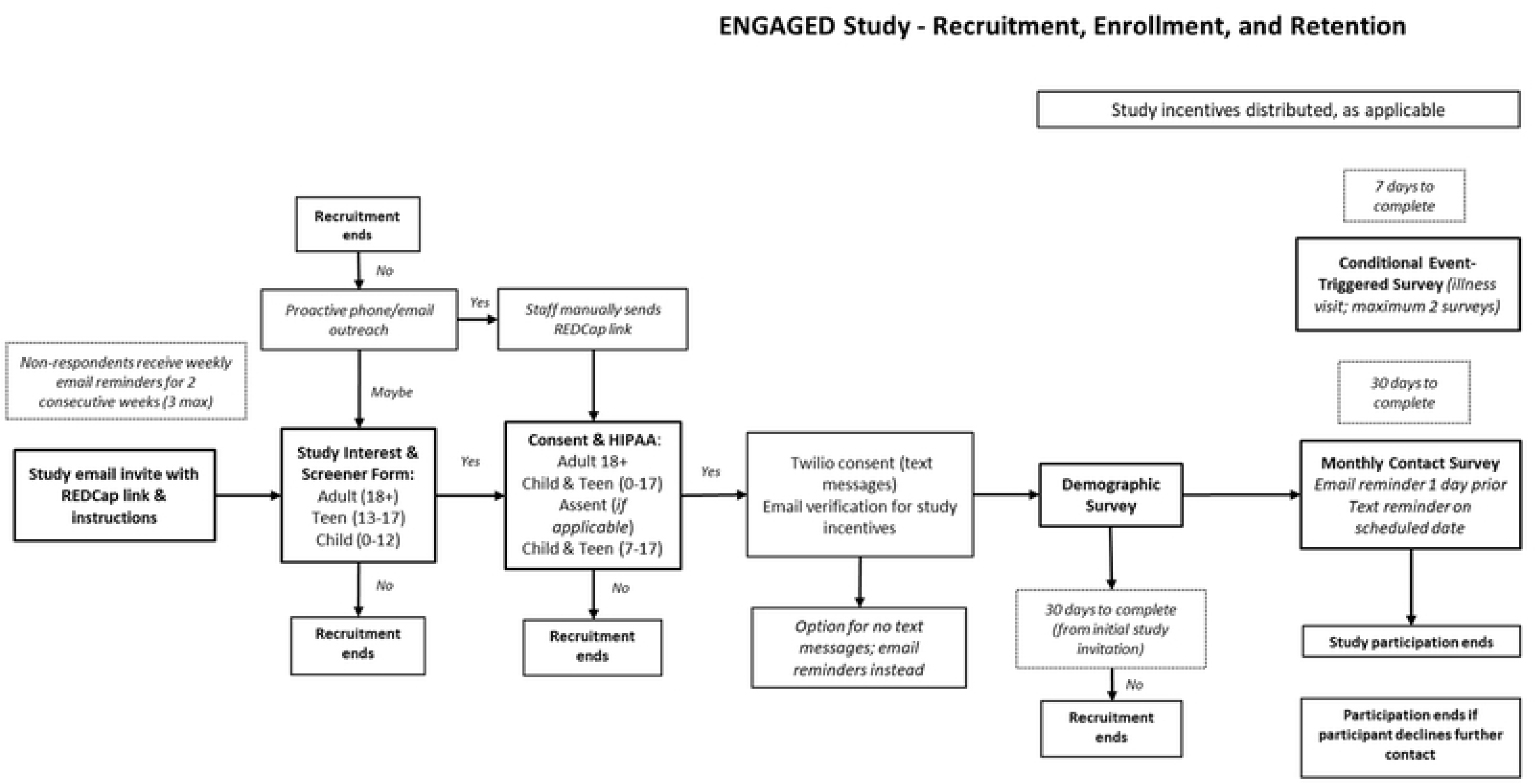
Study recruitment, enrollment, and retention steps.

Potential participants first completed an interest form, where they indicated definitive interest (opt-in), requested more information, or declined to participate (opt-out). Those who opted-in were directed to submit a brief screener to self-report if the participant and/or the reporting proxy met inclusion/exclusion criteria. If eligible, they were directed to a REDCap page where they (or their legal guardian) could provide informed consent and HIPAA authorization to enroll in the study, and permission to receive text message reminders for future study activities. All participants under age 18 were required to have a parent/guardian provide consent and HIPAA authorization on their behalf to enroll in the study. Participants aged 7-17 years also provided assent in addition to their parent/guardian’s consent for study participation.

When participants turned 7 or 18 years old during the study, they were sent assent and consent forms respectively. Each form was sent on the participants’ birthday and every following 3-days for non-respondents for a total of 3 invitations. Completion of these forms was required for continued study participation. Those who did not complete the appropriate assent/consent documents after 3 invitations were removed from future study contact, and their study data were censored on their birthday.

Once enrolled, study activities were fully automated and online. Additional live phone support was available to potential and enrolled participants as needed to answer questions about the study or study procedures.

### Study re-consent

In April 2025, a vendor change occurred for study incentives that required all actively enrolled study participants to re-consent to continue participating in the study.

After IRB approval, actively enrolled participants received an email invitation informing them of the incentive vendor change with instructions and an online link to the secure REDCap study portal to provide an updated consent or parental permission (and assent as applicable) to an IRB-approved consent addendum explaining the vendor change and participants’ new vendor options. Participants had the choice to stay with their current incentive vendor, which offered incentives to limited to one retailor (e.g. Amazon e-codes) or switch to the new study incentive vendor with multiple cash options to choose from multiple retailors. The main study consent was also updated for future participants to include the new study incentive vendor information.

## Data collection processes

### Demographic survey

After participants consented to participating in the study, they were immediately directed to a demographic survey. The demographic survey asked about participants’ age, gender, race, ethnicity, education, occupation, routine commute/travel patterns, study-specific baseline characteristics for vaccination status (influenza, COVID-19, respiratory syncytial virus), household income, and household size.

### Contact surveys

#### Monthly contact survey, self-managed symptoms

Two days after completing the demographic survey, participants received their first of six monthly contact surveys (Figure 2). A day prior to scheduled monthly survey distribution, participants received a pre-survey email reminder to help them reflect on contacts to share for the ‘day of interest’ when completing the contact survey. In addition, on the day that scheduled contact surveys were deployed, participants received a generic text message (for those who consented to receive text messages) or email reminder within 20 mins of the survey email invitation to remind them to check their email for the survey invitation and complete the survey. Participants who did not complete their monthly contact survey received additional weekly email reminders for 2 consecutive weeks to encourage survey response completion. This process was repeated each consecutive month for a total of 6 monthly contact surveys.

**Figure 2:**
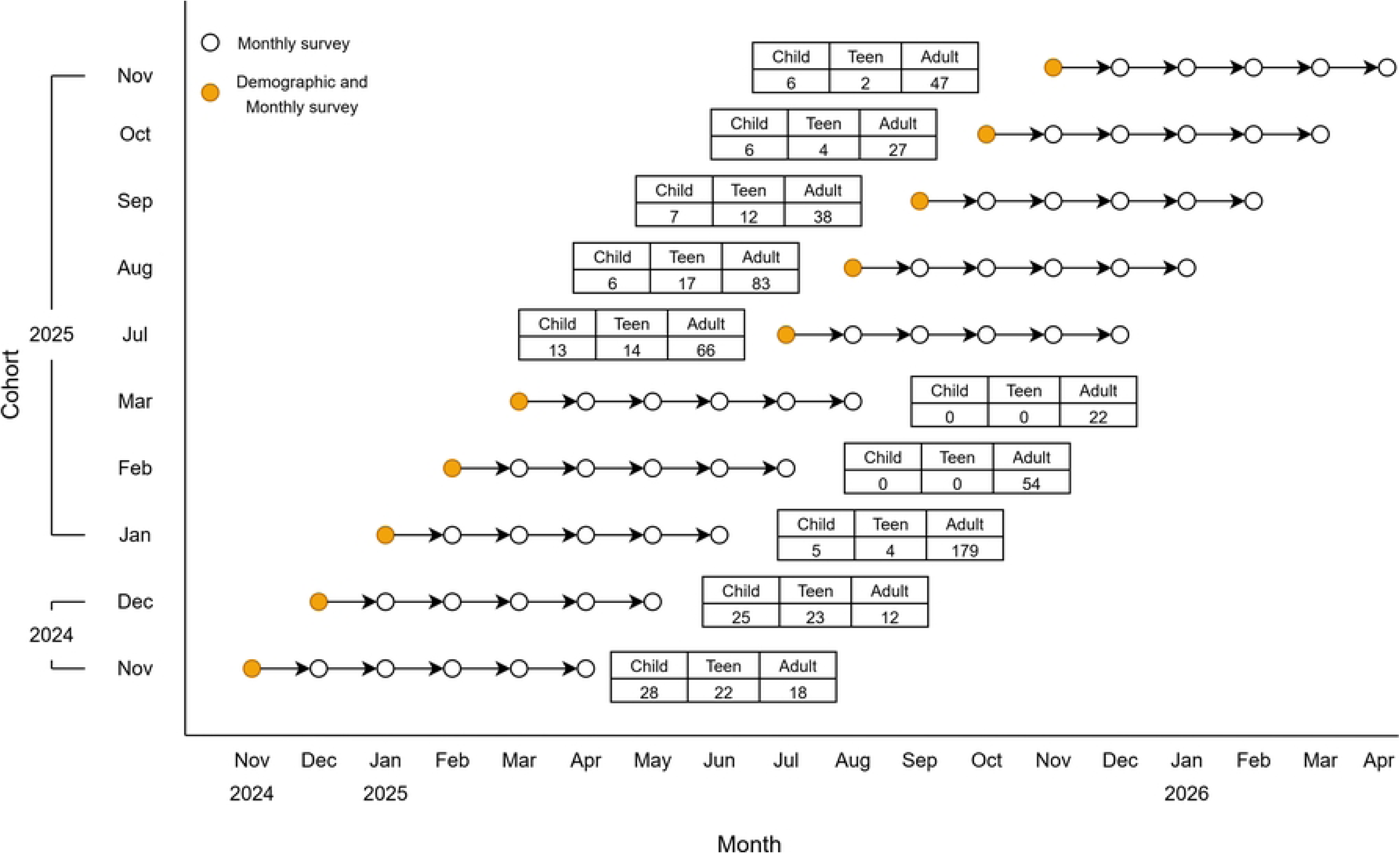
Monthly participant enrollment by cohort and survey completion timeline.

At the beginning of each monthly contact survey, participants were asked whether they had any symptoms related to acute gastroenteritis (AGE) or acute respiratory infection (ARI) in the past seven days, such as coughing, sore throat, sinus congestion, runny nose, fever, diarrhea, or vomiting. They were also asked to specify the date symptoms started and when they felt the sickest. If the date participants felt most sick fell within seven days of the date the survey was filled out, participants were asked to report on their contacts the day they felt most ill. If participants did not report AGE/ARI symptoms or if the date participants felt sickest was not within seven days of the date of survey completion, participants would instead provide information on their contacts for the prior day to completing the survey. (See supplemental materials)

#### Event-triggered contact survey, medically-attended symptoms

We conducted twice-weekly queries of participants’ electronic medical records for identification of medically-attended AGE and ARI with a lookback period of seven days. Additional event-triggered surveys were sent if a participant’s queried medical record showed a recent ambulatory, urgent care, and/or telehealth visit with ICD-10 diagnosis code(s) associated with AGE or ARI (see Supplemental form 9). Participants could receive up to two additional event-triggered contact surveys throughout their 6 months of active study participation.

At the beginning of each event-triggered contact survey, participants were asked to report their symptoms, symptom start date, and the date they felt the sickest. If the date participants felt the sickest fell within 7 days of the date the surveys were opened, participants were asked to report on their contacts the day they felt most ill. Otherwise, the survey ended if reported ‘most ill’ date fell outside the 7-day reporting window.

#### Contact data collected

In each of the contact surveys, participants were asked about their interactions on the day of interest specified in the survey (either the day prior or, if symptomatic, the day they felt most sick if within 7 days of the survey).

We asked participants to report each interaction in one of three categories: individual contacts, other one-on-one contacts, and group contacts (see Table 1). Individual contacts were contacts with people whom the participant could name individually such as household members, friends, and work colleagues. We defined individual contacts as a two-way (one-way for young children not yet speaking) conversation with five or more words in the physical presence of another person or physical skin-to-skin contact. Individual contacts were further broken down into household and non-household contacts. Household contacts were defined as contacts with people who lived in the same residence in which participants regularly slept, such as family members or roommates. For the household contacts, we asked participants to report number of contacts, age group (within 5-10 years) and gender of the contacted person, duration, and whether it involved physical contact. For the non-household contacts, in addition to all the variables in the household contacts, we asked location, indoors/outdoors, and typical frequency of contacts (e.g. daily, weekly).

**Table 1:** Summary of the definition and data collected for each contact type.

| Type of Contact | Individual contacts | Other one-on-one contacts | Group contacts |
| --- | --- | --- | --- |
| Contact definition | One-on-one in-person interactions with people you can name individually<br>- a two-way (one-way for young children not yet speaking)<br>conversation with five or more words in the physical presence of another person, or<br>- physical skin-to-skin contact (for example a handshake, hug, kiss,<br>or contact sports) | One-on-one in-person interactions with people you cannot name individually e.g. serving customers at cash register | Group of people with whom you talked, interacted, or shared space with e.g. in a meeting/classroom, public transit, in a doctor's office waiting room, athletic event, church service |
| Data collected | Household member or not<br>Age<br>Gender<br>Duration<br>Physical contact<br>Location<br>Indoor/outdoor<br>Frequency | Number of contacts<br>Average age category | Approx. number of people in group<br>Indoor/outdoor<br>Duration<br>Average age category |

Other one-on-one contacts were contacts with people at school or a workplace who the participants could not individually name, such as seeing customers at a checkout. Participants were asked the approximate number of one-on-one contacts they had and age groups of those people (adults, teenagers/children). Group contacts were contacts with a group of people with whom participants talked, interacted, or shared space such as attending a meeting or being on public transit. Participants were asked about the location, duration, setting (indoors/outdoors), age groups, and approximate number of people in each group setting.

### Study incentives

Each enrolled study participant received up to $260 in study compensation for completing up to 8 contact surveys and the demographic survey. Incentives for completing study surveys were divided as follows - $20 for demographics, $20 each for months 1 and 2 contact surveys, $30 each for months 3 and 4, and $40 each for months 5 and 6. Participants received $30 for each of the additional event-triggered surveys.

XI-axis refers to a calendar month and y-axis refers to the months each cohort were recruited. The circle indicates the months contact surveys were sent to participants in each cohort. The number of participants recruited in each cohort by age groups (Child: <13 years old, Teen: 13-17 years old, Adults: >18 years old) are shown in the table. The age distribution of each cohort varied throughout the study as recruitment efforts were adjusted in response to age-specific differences in enrollment. This was necessary to meet overall age-specific recruitment targets across seasons. Not pictured is up to two additional event-triggered surveys per participant who sought care at KPGA facilities for AGE/ARI symptoms (event-triggered surveys). New recruitment was paused in April, May, and June of 2025 in response to changes in federal funding, but data collection continued as usual for participants already enrolled.

### Quantitative data analysis

#### Number of contacts

The number of contacts per participant will be defined as a) the number of individual contacts (i.e. number of people with 1-on-1 interaction), b) the number of group contacts (i.e. number of groups), and c) the total number of contacts (i.e. number of people reported as individual contacts plus the number of people reported from groups). Group contacts will be incorporated into a total number of contacts and overall contact matrices. Number of contacts reported in group contacts will be approximated using the midpoint of the category, and scaled by contact duration. Age distributions of group contacts will be imputed based on reported age categories of a group, participant age, and location, to reflect the context of participants’ reported contact settings.

#### Age-specific contact matrices

We will produce contact matrices of the mean number of daily contacts between age groups. At least four sets of contact matrices will be produced: a) <u>overall</u> for all monthly contact surveys, b) among <u>healthy</u> people reporting no recent AGE/ARI symptoms, c) among people with <u>self-managed</u> AGE/ARI symptoms (that did not progress to medically-attended symptoms), and d) among those with <u>medically attended</u> AGE/ARI illness who sought care at KPGA. These matrices will be adjusted for population demographics (age, gender, and race) and will account for reciprocity as necessary.

#### Characterizing social contacts

We will estimate the total number of contacts, number of individual contacts, and number of groups contacts per participant by participant characteristics, including age, gender, household membership, and school/work participation. We will report mean, median, and interquartile (IQR) of number of contacts. We will also stratify contacts by duration of contact and context (i.e. whether the contact was indoors or outside, at home or at work/school). Monthly contact surveys and event-triggered surveys will be evaluated for duplicate illnesses for self-managed illness that progressed to medically-attended illness. We will calculate and compare contact patterns by season (e.g. school breaks, holidays, summer/winter), participants’ illness status (e.g. well, self-managed symptoms, medically-attended symptoms), and day of the week.

#### Predictors and effects of number and type of contacts

We will perform regression analysis to assess associations between participant characteristics and number and type of contacts (one-on-one versus group contacts, home versus work, indoor versus outdoor, etc.). Random effects or similar methods will be used to account for repeated measurements from the same individuals. We will calculate the average number of contacts by type, and the distribution of contact age groups by participant age group.

### Dissemination plans

This study collected data from a population in Georgia as a part of the InsightNet group, a multi-site collaboration with the effort to collect data across US and potentially compare the contact patterns across regions. Data will be harmonized and reported alongside other studies from the InsightNet group. Upon study completion, de-identified individual level contact data as well as contact matrices and analytical code will be made publicly available. Data structures will follow the format of data standards used in socialcontactdata.org (15) which include participant, contact, survey day, and dictionary datasets. We will publish our data on Zenodo platform for public use, and scripts used for the data analysis such as creating contact matrix will be made available through GitHub/Zenodo platform upon analysis completion. Study findings will be published in scientific journals and presented at conferences. Furthermore, we currently have a publicly available dashboard to share study progress and key data visualizations (<u>ENGAGED</u> <u>dashboard</u>).

## Discussion

To our knowledge, this study is the first study collecting social contact data on a population level in metropolitan Atlanta area in Georgia, US with six months of repeated data collection per participant to capture social behavior change by seasonality and symptom status.

Understanding the contact characteristics of people when they are feeling unwell or seeking healthcare intervention is crucial, as we can incorporate those changes into ID transmission models. We also collected multiple types of contacts (group contacts and one-on-one contacts) in addition to individual contacts, which will add value by presenting a more complete picture of social behavior and disease transmission potential.

We also acknowledge some limitations. First, this study recruited participants mostly in urban or suburban areas of Georgia which may have different social contact patterns than rural areas, limiting the transportability of data to non-urban settings. Secondly, participants may preferentially report contacts of longer duration or with individuals who they frequently interact with. This study asked participants to fill in their contacts on the day of interest (previous day of the survey or the day they felt most sick within 7 days of survey); therefore, responses depended on participants’ ability to accurately recall contacts for that day. Lastly, because contact surveys for minors were completed by their caregivers, they might not reflect the actual contacts and may potentially under- or overestimate the number or duration of contacts.

However, this study adds a value to existing social mixing data by providing longitudinal contact survey data and assessing individual-level behavioral change when participants are ill or by season.

## Supplementary Materials

See supplementary materials document.

## Data Availability

Upon study completion, de-identified individual level contact data as well as contact matrices and analytical code will be made publicly available.

## ACKNOWLEDGEMENTS

We thank Lala Zewdu and Joe M. Chery for their continuous support in study implementation.

Lala (KP): BS; Center for Research and Evaluation, Kaiser Permanente Georgia, Atlanta, GA, USA Joe (KP): BS, Center for Research and Evaluation, Kaiser Permanente Georgia, Atlanta, GA, USA

